# Longer-Term Geospatial Food Access and the Incidence of Breast Cancer in Metropolitan Chicago

**DOI:** 10.1101/2025.09.11.25335064

**Authors:** Niyati Sudhalkar, Neng Wan, Vanessa M. Oddo, Caryn E. Peterson, Jiehuan Sun, Garth Rauscher

**Affiliations:** School of Public Health, Division of Epidemiology and Biostatistics, University of Illinois Chicago; School of Environment, Society, and Sustainability, University of Utah; Department of Kinesiology and Nutrition, College of Applied Health Sciences, University of Illinois, Chicago; University of Illinois, Chicago Center for Global Health; University of Illinois Cancer Center

**Keywords:** breast cancer incidence, residential history, geospatial food access, inverse distance weighting, nearest distance measures, neighborhood food environment

## Abstract

Diet quality contributes to breast cancer (BC) risk and is shaped in part by the neighborhood food environment. Yet, the long-term impact of residential food environments on BC incidence remains largely underexplored. We linked residential histories of 7,396 BC cases and 21,900 controls in Chicago (1990–2019) to food outlet data from the National Establishment Time Series. Cumulative (time-weighted), inverse distance-weighted (IDW), food access scores were derived along with nearest distance metrics for “*healthy*” and “*less healthy”* food access. Associations with incidence were stronger for walking than driving distance-based measures and for nearest distance-based measures over IDW measures. In multivariable logistic regression models, BC incidence decreased monotonically with shorter walking distance to the nearest healthy food outlet, reaching a 60% lower incidence for shorter vs. longer walking distances (OR=0.42, 95% CI=0.38, 0.48). Similarly, incidence increased monotonically with shorter walking distance to the nearest *less* healthy food outlet, reaching a 150% greater incidence for shorter vs. longer walking distances (OR=2.49, 95% CI=2.19, 2.83). This is the first study to use residential histories to define long-term, time-weighted geospatial food access metrics in BC epidemiology, highlighting how cumulative neighborhood environments have the potential to shape cancer risk and informing targeted interventions.

## BACKGROUND

Breast cancer is the second most common cancer among women in the United States, affecting approximately 1 in 8 women during their lifetime, with incidence highest in Non-Hispanic (NH) White vs. NH Black women.^1–3^ About one-third of breast cancer cases have been attributed to preventable factors, including an unhealthy diet and obesity, among other lifestyle factors. A western dietary pattern, characterized by high consumption of calorie-dense foods, red meat, processed foods, and low consumption of fruits and vegetables are pro-inflammatory and contribute to obesity,^4,5^ which, in turn, leads to hormonal imbalances that could increase risk of hormone receptor-positive breast cancer,^6,7^ particularly in postmenopausal women.^8–10^ In addition, a *less* healthy diet has been linked to a proinflammatory environment that might lead to a greater incidence of estrogen and progesterone receptor (ER/PR) negative breast cancer.^11,12^ Conversely, healthier dietary patterns help maintain hormonal balance and reduce breast cancer risk, with protective effects seen especially for premenopausal women and for both hormone receptor-positive and hormone receptor-negative tumors.^11^

Potential neighborhood-level risk factors include access to healthy food, physical and chemical exposures in the environment, and socioeconomic resources (e.g., area-level deprivation), which affect breast cancer risk through their impacts on biological stress, social support, and dietary patterns in an individual.^13–16^ Residential proximity to food outlets in the neighborhood influences individual-level food purchasing behaviors and diet quality.^17–20^ Poor food environments – characterized by limited access to fresh produce and high-quality groceries are associated with *less* healthy dietary patterns.^21^ Limited access to healthy foods can result in nutritional deficiencies and associated adverse health outcomes, such as colorectal cancer,^2,22,23^ even after adjusting for socio-economic factors.^2^

Nevertheless, the association between neighborhood food environment and breast cancer incidence largely remains unexplored. Moreover, most studies of the food environment and other health outcomes have relied on residence at diagnosis as a proxy for neighborhood exposures since long-term residential histories are difficult to capture.^15,24^ However, the emerging availability of commercial sources like Lexis-Nexis for linking patients to their residential histories over many years has facilitated the study of longer-term food access.^25,26^ Long-term residential histories allow for a comprehensive evaluation of the cumulative impact of neighborhood exposures such as the food environment.

For this study, we analyzed up to 30 years of residential histories (1990–2019) for women seeking breast health care from a large healthcare organization in the greater Chicago area. We calculated walking and driving distance-based measures of geospatial access to healthy and *less* healthy food, examined associations with breast cancer incidence, and explored differences by tumor subtype and menopausal status. We hypothesized that greater access to healthy food in the residential environment would be associated with reduced breast cancer incidence, and vice versa.

## METHODS

### Study Population

Data came from a previously constructed case-control study of women receiving breast screening and diagnostic imaging and biopsy procedures between 2002 and 2018 from a single large health care organization. The study was approved by the Institutional Review Boards (IRBs) at Advocate Health Care, the University of Illinois, Chicago (UIC), and the Illinois

Department of Public Health (IDPH).^27,28^ Roughly three controls (n=23,172) were frequency-matched to each case (n=7,682) on year of entry into the cohort (year of first exam), total duration in the cohort (year of most recent exam minus year of first exam), and 5-year age group. We submitted patient identifiers to Lexis-Nexis for linkage to obtain residential histories (RH).

All cases and controls had at least one linked address: 82% (86,453 of 105,080) of linked addresses were within the catchment area, and 95% (82,286 of 86,453) of linked addresses within the catchment area were from the urban portion of the catchment area (Table 1). Food access measures were constructed based on 18,990 addresses for 7,396 cases and 62,972 addresses for 21,900 controls.

**Table 1.** Distribution of Cases and Controls and their residential history addresses, in Breast Cancer Case-Control Study (1990-2019)

|  | **Total**<br>**(N)** | **Cases**<br>**(n1)** | **Controls**<br>**(n2)** |
| --- | --- | --- | --- |
| Case-control Parent Study | 30,854 | 7,682 | 23,172 |
| Submitted identifiers for RH linkage | 30,854 | 7,682 | 23,172 |
| Individuals with at least one linked address | 30,854 | 7,682 | 23,172 |
| Geocoded addresses | 105,080 | 23,197 | 81,883 |
| Geocoded addresses within the catchment area | 86,453 | 20,008 | 66,445 |
| Individuals within the catchment area | 30,038 | 7,632 | 22,406 |
| Geocoded addresses within the Urban catchment area | 82,286 | 19,155 | 63,131 |
| Individuals within the Urban catchment area | 29,395 | 7,487 | 21,908 |
| Addresses with Food access scores<sup>a</sup> | 81,962 | 18990 | 62,972 |
| Individuals with Food access scores<sup>a</sup> | 29,296 | 7,396 | 21,900 |

<sup>a</sup> Cumulative Inverse Distance Weighted, Time-weighted food access scores computed for each case and control for walking (within 1.5 miles) and driving (within 5 miles) distances from residence to healthy vs. *less* healthy food outlets.

### Catchment area of analyses

We purchased annual data (1990-2021) on locations and types of food outlets from the National Establishments Time Series (NETS) for Combined Statistical Area (CSA) FIPS 176- Chicago Naperville, IL-IN-WI 2019; CSA-176 (2019), covering 19 counties across Illinois, Indiana, and Wisconsin. This CSA served as the total catchment area for these analyses. Since more than 90% of the residences used in this study were urban or suburban (Figure S1), we restricted analyses to urban and suburban patient residences within the CSA.

### Food outlets

From NETS we extracted information for each food outlet including spatial identifiers (latitude, longitude, street address, and FIPS codes), tradename, company name, address, 8-digit Standard Industrial Classification (SIC) code, establishment type (public/private), and food outlet/industry type (e.g. deli, grocery store, supermarket, restaurants, etc.). With guidance from prior studies that classified outlets,^29–31^ we classified food outlets as healthy or *less* healthy based on SIC codes, industry type, and trademarks. Healthy outlets included grocery stores, supermarkets, and fresh fruits and vegetables markets, while *less* healthy outlets included convenience stores, gas stations with convenience stores, and fast-food places. Cafés, restaurants, delis, sandwich shops, and bars were categorized as undetermined and excluded from our analyses (Table S1).

### Measuring Geospatial Access

The ESRI network analysis tool, Origin-Destination Cost Matrix (ODCM), was used to compute travel distances in ArcGIS Pro 3.2.3 from each patient residence to each food outlet. Street network data is the foundation of network analysis as it provides information on edges and nodes used to identify intersections, streets, and types of roads (primary, secondary, highway, etc.) that are needed to measure distances between any two points. Since the census street files used to define street networks change with each decade, we created a street network for each decade of residential history from 1990 (to calculate distances for years 1990-1995), 2000 (for years 1996-2005), 2010 (2006-2015) and 2019 (2016-2019).

#### Inverse distance-based measures

We used ESRI’s Model Builder in ArcGIS Pro to automate an OCDM analysis for walking and driving distances from each patient residence (origins) to food outlets (destinations) over all years of food outlet data (1990-2019). We excluded residential histories occurring after the year of diagnosis for cases and after the last year of clinical data for controls. For each patient-year of residential history, we calculated walking distances from the patient’s address to each healthy food outlet within 1.5 miles of their residence. This threshold reflects common use in prior food environment and built environment research as a reasonable upper bound for walkable access in urban settings, often corresponding to a 30-minute round-trip walk.^32,33^ The inverse distances to each food outlet were summed to create a healthy food access score for walking. Then, for each patient-year of residential history, we calculated driving distances from the patient’s address to each healthy food outlet within 5 miles of their residence and summed the inverse driving distances to create a healthy food access score for driving. This threshold is frequently used in the literature as a meaningful catchment area for driving access to food retail, particularly in urban and suburban contexts.^32,34^. For both walking and driving distance-based scores, we assumed a linear decay such that the importance or relevance of a food outlet diminished in a linear fashion with distance from each patient’s residence. Then, the annual distance-based healthy food access scores for each patient were converted to a time-weighted average score, which equates to a simple average of all available years of data. We repeated an identical process for defining time-weighted, inverse distance-weighted (IDW) scores for walking and driving distances to *less* healthy food outlets. Finally, we defined variables for the ratio of healthy to unhealthy food access scores, separately for driving and walking distance-based measures.

#### Nearest distance-based measures

For each patient-year of residential history, we computed the nearest walking (and driving) distance to healthy (and *less* healthy) food outlets. These annual nearest distance measures were converted to time-weighted average scores for walking (and driving) distance to healthy (and *less* healthy) food outlets.

All data management and analyses were conducted using ArcGIS Pro 3.2.3 and STATA 18.5.

### Sociodemographic, Reproductive, Food Access, and Other Variables

The final dataset for this case-control study consisted of 7,396 cases and 21,900 controls. Reproductive factors included age at menarche, menopausal status, parity and number of live births, breastfeeding in months, and late parity (defined as a live birth after age 35). In addition, the dataset included variables for first-degree family history of breast cancer (none, second-degree only, first-degree, and multiple first-degree or early onset before age 50), and any history of breast biopsy. For sub-analyses, cases and controls were stratified by race and by menopausal status, and breast cancer cases were sub-grouped into ER/PR-negative and ER/PR-positive tumors.

### Statistical Analysis

We constructed a Directed Acyclic graph (DAG) (Figure S2) to specify our assumed causal relations between pre-diagnostic food access scores, breast cancer incidence, and *a-priori* risk factors. Summary statistics (mean, median, interquartile range, and standard deviation) were calculated, and histograms were used to characterize the distribution of food access scores and identify potential outliers. Spearman correlations were used to assess correlations between food access scores. Descriptive analyses compared food access scores for walking and driving by case and control status and sociodemographic, reproductive, and lifestyle-related factors.

To allow for non-monotonicity of associations and to facilitate interpretation, we categorized all food access measures into five groups at the quintiles. We then ran logistic regression models for the mutually adjusted associations for walking distance-based healthy and *less* healthy food access with breast cancer incidence (separately for IDW and nearest distance-based measures). We also ran logistic regression models for the mutually adjusted associations for driving distance-based healthy and *less* healthy food access with breast cancer incidence (separately for IDW and nearest distance-based measures). We also ran a logistic regression model for the ratio of healthy to unhealthy food access scores, and for the ratio of nearest distance to healthy vs. unhealthy food outlets. All models were adjusted *a-priori* for matching factors (age and year of entry and exit from the study), race/ethnicity, reproductive factors (menopausal status, breastfeeding, number of live births, age at menarche, and parity), family history of breast cancer, history of prior biopsy, as well as residential tract-level measures of disadvantage and affluence and tract-level racial/ethnic composition (proportion NH White, NH Black, and Hispanic.^16,35^

## RESULTS

### Summary statistics and distribution, and correlation of food access scores

Table S2 describes the summary statistics, and Figure S3 shows the distribution of all the food access scores for walking and driving on an absolute and relative scale (IDW and nearest distance-based measures), respectively. All food access scores for walking and driving on an absolute and relative scale (ratio measures) had a right-skewed distribution with high kurtosis, indicating outliers/extreme values. Correlations between IDW and nearest distance food access measures are shown in Figure 1A and 1 B, respectively. Overall, we observed that the distribution of all IDW-based food access scores by quintiles was comparable between cases and controls (Table S3), as was the distribution of food access scores by quintiles for nearest distance-based measures was comparable between cases and controls, while the proportion of cases were higher in the first quintile of healthy food and ratio measures and the fifth quintile of *less* healthy food (Table S4).

**Figure 1.**
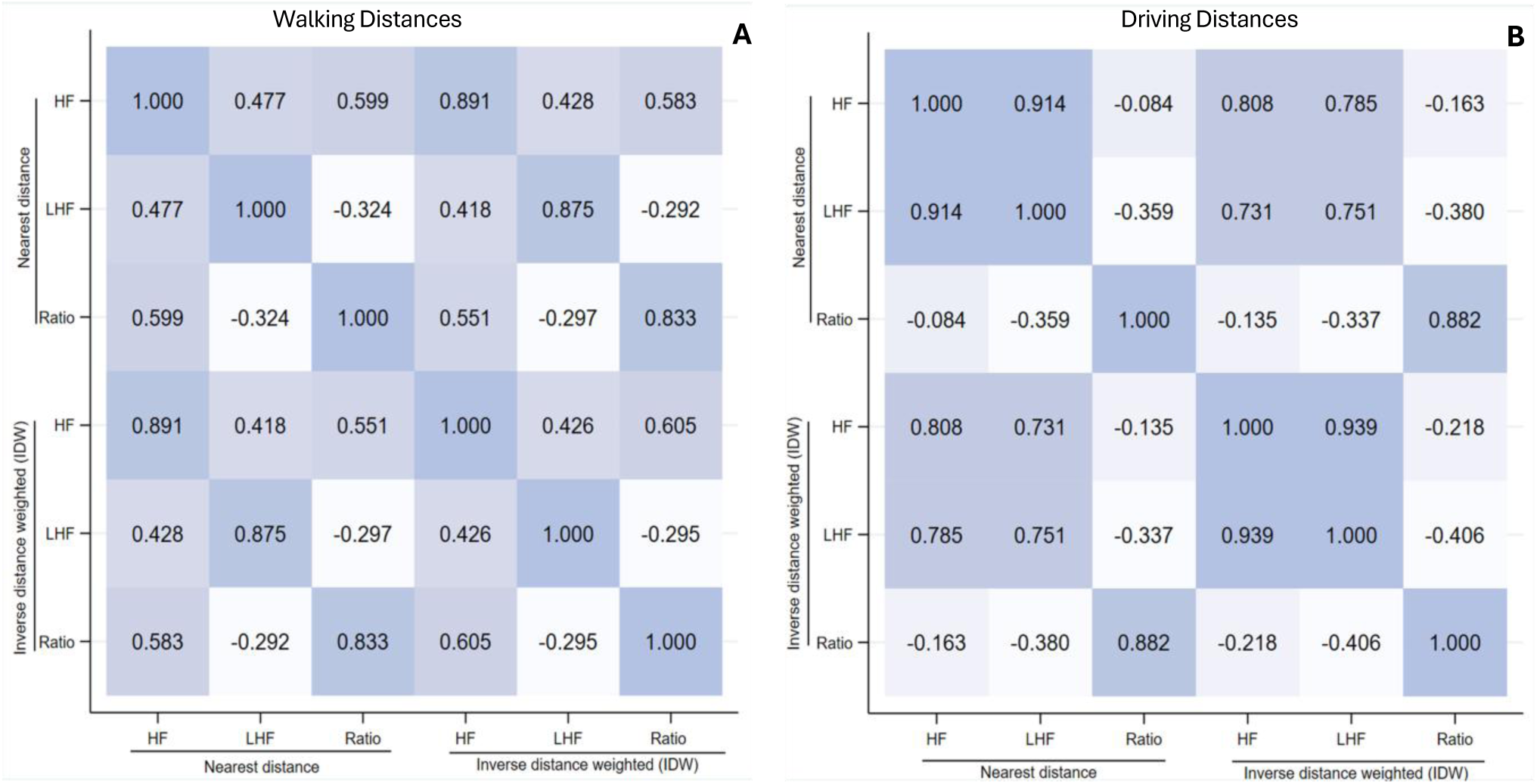
Correlation matrix for all food access scores (Nearest distance and IDW-based measures) for walking distances (A) and driving distances (B). Stronger correlations are shown using darker highlights, while lighter highlights show weaker correlations.

### Distribution of individual-level sociodemographic, reproductive and other risk factors

The distribution of sociodemographic factors, reproductive factors, and other risk factors is shown in Table 2. Pre-dominantly, 72% of cases and 55% of controls were of NH White race/ethnicity. Cases were more likely to be post-menopausal, with a family history of breast cancer, have no live births, and no prior history of biopsy, as compared to controls. The distribution of age at menarche, smoking and BMI was comparable between cases and controls (Table 2). Later age at menarche (at or above age 15), parity (one or more livebirths), early parity (before age 35), and more than six months of breastfeeding were each associated with lower breast cancer incidence. Increased breast cancer incidence was associated with a family history of breast cancer, a history of biopsy, post-menopausal status, and late parity. Cases belonged to neighborhoods with marginally higher affluence and lower disadvantage, whereas the racial/ethnic composition of the neighborhoods at the tract level was comparable between cases and controls.

**Table 2:** Crude Distribution of Sociodemographic, reproductive, lifestyle-related, and other risk factors in breast cancer case-control study.

|  | Case (n=7396) |  | Control (n=21,900) |  |
| --- | --- | --- | --- | --- |
|  | n | % | n | % |
| Race / Ethnicity |  |  |  |  |
| NH White | 5314 | 72% | 11,966 | 55% |
| NH Black | 1642 | 22% | 5613 | 26% |
| Hispanic | 391 | 5% | 1846 | 8% |
| Other | 49 | 1% | 2475 | 11% |
| Family history of BC (1st degree) |  |  |  |  |
| None | 5302 | 74% | 17,251 | 84% |
| Any | 1230 | 17% | 2,501 | 12% |
| Multiple/Early Onset | 626 | 9% | 832 | 4% |
| Any prior history of biopsy |  |  |  |  |
| No | 5,887 | 82% | 17,515 | 89% |
| Yes | 1,254 | 18% | 2,193 | 11% |
| Ever Smoker |  |  |  |  |
| No | 3,925 | 69% | 10561 | 68% |
| Yes | 1,730 | 31% | 4,888 | 32% |
| Body Mass Index (BMI) |  |  |  |  |
| Underweight (<18.5 kg/m<sup>2</sup>) | 168 | 2% | 507 | 2% |
| Normal (18.5- 24.9 kg/m<sup>2</sup>) | 1,247 | 17% | 3,674 | 17% |
| Overweight (25- 29.9 kg/m<sup>2</sup>) | 1,659 | 22% | 4,892 | 22% |
| Obese (Class I) (30-34.99 kg/m<sup>2</sup>) | 1,176 | 16% | 3,486 | 16% |
| Obese (Class II-III) (>35 kg/m<sup>2</sup>) | 3,146 | 43% | 9,341 | 43% |
| Post-Menopausal |  |  |  |  |
| Yes | 6,181 | 84% | 15,907 | 73% |
| No | 1,215 | 16% | 5,993 | 27% |
| Age at menarche |  |  |  |  |
| < 11 years | 1,293 | 20% | 3,432 | 20% |
| 12-14 years | 4,494 | 69% | 11,593 | 67% |
| >15 years | 753 | 12% | 2,397 | 14% |
| Parity |  |  |  |  |
| Nulliparous | 1,641 | 22% | 4,370 | 22% |
| Early (<35 years) | 5,368 | 73% | 14,592 | 74% |
| Late (>35 years) | 387 | 5% | 897 | 5% |
| Number of live births |  |  |  |  |
| 0 | 1,524 | 21% | 3035 | 15% |
| 1 | 1,033 | 14% | 3,023 | 15% |
| 2 | 2,237 | 31% | 6288 | 31% |
| 3 | 1,480 | 20% | 4411 | 22% |
| ≥4 | 1,058 | 14% | 3538 | 17% |
| Breastfeeding (months) |  |  |  |  |
| 0 months | 4,670 | 63% | 13,589 | 62% |
| 1 to 6 months | 2,703 | 37% | 8,158 | 37% |
| 7 to 18 months | 23 | 0.30% | 153 | 1% |

### Geospatial access to food outlets and the overall risk of breast cancer

#### Healthy food access

*Walking distance:* Greater walking distance-based IDW scores for healthy food access demonstrated a U-shaped association with breast cancer incidence, with ∼30% lower incidence in the second, third and fourth quintiles and returning to an incidence in the highest fifth quintile similar to that for the lowest quintile (Table 3 and Figure 2). Associations were stronger for walking distance to the *nearest* healthy food outlet, with incidence dropping monotonically to a 60% reduced incidence in the highest vs. the lowest quintile compared to the lowest fifth (Table 4 and Figure 3).

**Figure 2.**
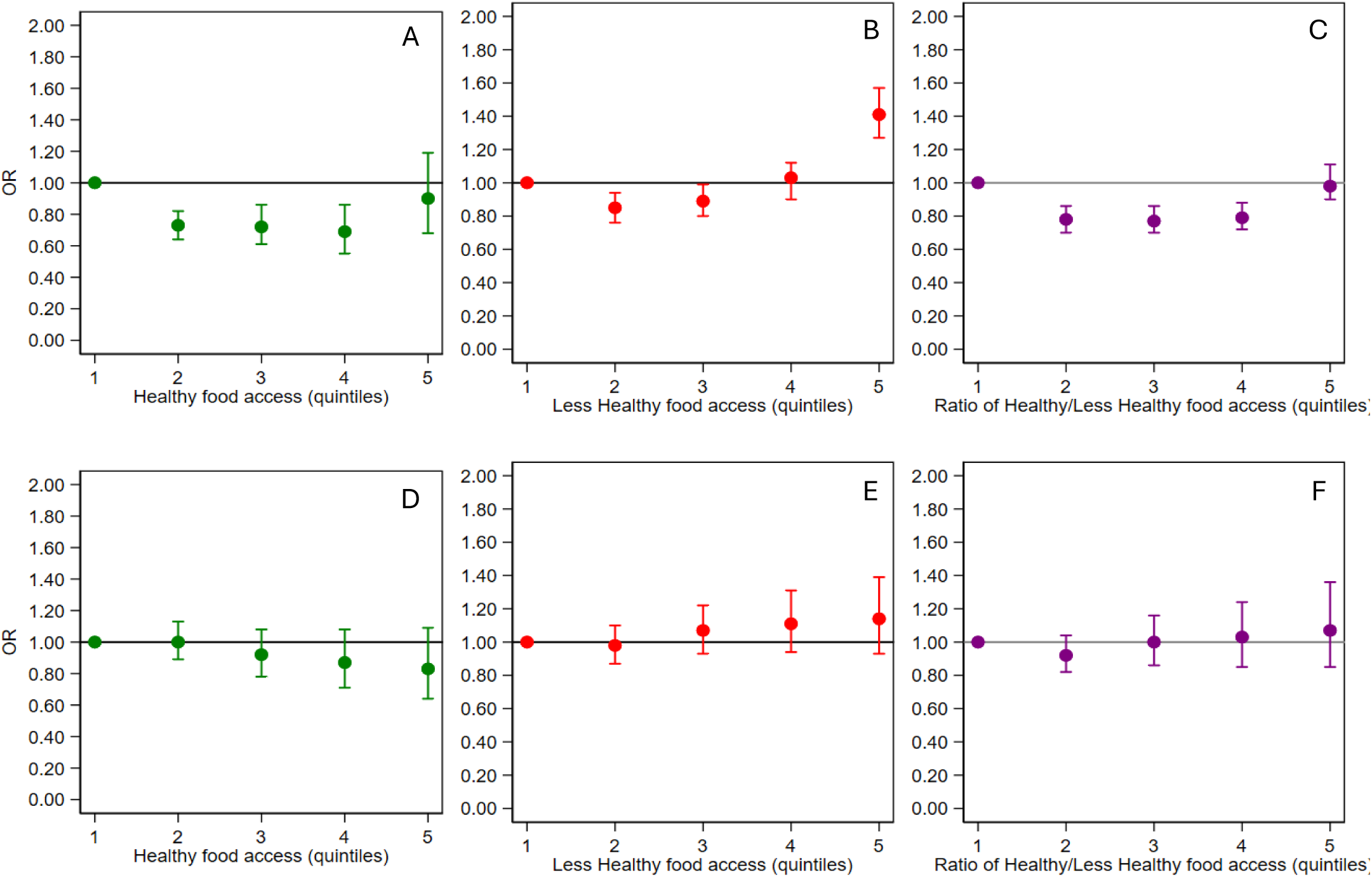
Mutually adjusted associations with breast cancer incidence for Inverse-Distance Weighted (IDW) healthy food access scores (green), *less* healthy food access scores (red), and the ratio of healthy to *less* healthy food access scores (purple). Walking distance-based measures are shown in the top row; driving distance-based measures are shown in the bottom row.

**Figure 3.**
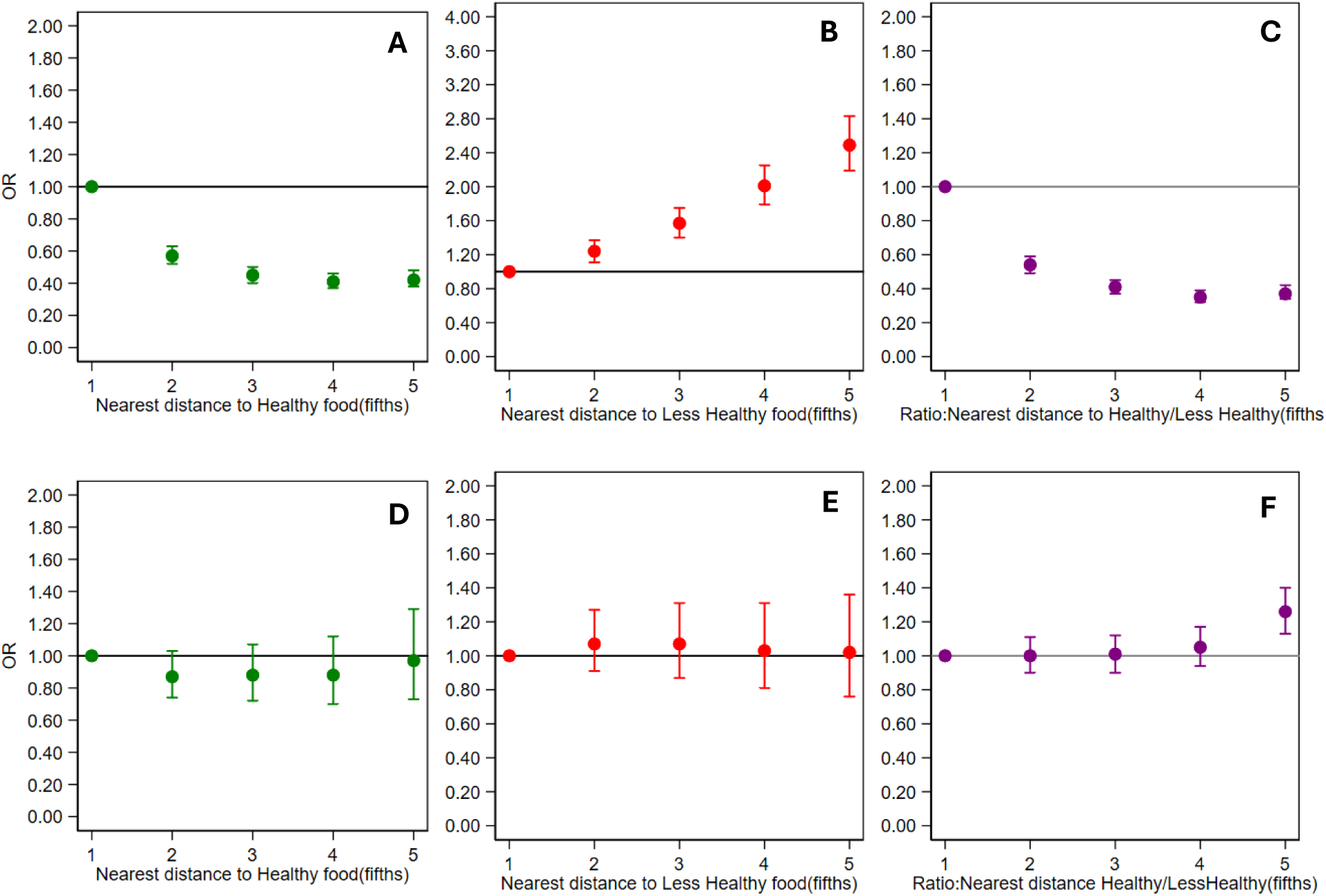
Mutually adjusted associations with breast cancer incidence for distance to *nearest* healthy food outlet (green), less healthy food outlet (red), and the ratio of distances to nearest healthy and less healthy food outlet (purple). Walking distance-based measures are shown in the top row; driving distance-based measures are shown in the bottom row.

**Table 3.** Associations with breast cancer incidence for mutually adjusted IDW healthy and *less* healthy food access scores (and their ratio), separately for walking and driving distance-based <u>measures.</u>

|  | Walking |  |  | Driving |  |
| --- | --- | --- | --- | --- | --- |
|  | OR<sup>a</sup> | 95% CI |  | OR<sup>a</sup> | 95% CI |
| Healthy (fifths/quintiles) |  |  |  |  |  |
| First |  | Referent |  |  | Referent |
| Second | 0.73 | 0.64 - 0.82 | ** | 1.00 | 0.89 - 1.13 |
| Third | 0.72 | 0.61 - 0.86 | ** | 0.92 | 0.78 - 1.08 |
| Fourth | 0.69 | 0.55 - 0.86 | ** | 0.87 | 0.71 - 1.08 |
| Fifth | 0.90 | 0.68 - 1.19 |  | 0.83 | 0.64 - 1.09 |
| *Ordinal* | 0.97 | 0.93 - 1.07 |  | 0.95 | 0.89 - 1.01 |
| *Less* healthy (fifths/quintiles) |  |  |  |  |  |
| First |  | Referent |  |  | Referent |
| Second | 0.85 | 0.76 - 0.94 | ** | 0.98 | 0.87 - 1.10 |
| Third | 0.89 | 0.80 - 0.99 | * | 1.07 | 0.93 - 1.22 |
| Fourth | 1.00 | 0.90 - 1.12 |  | 1.11 | 0.94 - 1.31 |
| Fifth | 1.41 | 1.27 - 1.57 | ** | 1.14 | 0.93 - 1.39 |
| *Ordinal* | 1.10 | 1.08 - 1.13 | ** | 1.04 | 0.99 - 1.09 |
| Ratio (fifths/quintiles) |  |  |  |  |  |
| First |  | Referent |  |  | Referent |
| Second | 0.78 | 0.70 - 0.86 | ** | 0.92 | 0.82 - 1.04 |
| Third | 0.77 | 0.70 - 0.86 | ** | 1.00 | 0.86 - 1.16 |
| Fourth | 0.79 | 0.72 - 0.88 | ** | 1.03 | 0.85 - 1.24 |
| Fifth | 1.00 | 0.90 - 1.11 |  | 1.07 | 0.85 - 1.36 |
| *Ordinal* | 1.00 | 0.98 - 1.03 |  | 1.03 | 0.97 - 1.09 |

IDW = inverse distance weighting
\*\*\* p<0.0001, \*\* p<0.001, \* p<0.01 ^Cumulative Time weighted Food Access scores (computed using the Inverse Distance weighting method).
<sup>a</sup> Adjusted for age, year of entry, and exit from the study, individual-level race/ethnicity, reproductive factors (menopause, livebirths, breastfeeding, parity, age at menarche), tract-level SES factors (disadvantage, affluence, proportion of NH, White, Black, and Hispanic at tract level), family history of breast cancer and prior history of biopsy and, mutually adjusted for food access scores; models with healthy food scores are adjusted with scores for *less* healthy food and vice versa, except for the ratio measures that are implicitly adjusted.

**Table 4.** Associations with breast cancer incidence for mutually adjusted distance to nearest healthy and *less* healthy food outlets (and their ratio), separately for walking and driving distance- based measures.

|  | Walking |  |  | Driving |  |  |
| --- | --- | --- | --- | --- | --- | --- |
|  | OR<sup>a</sup> | 95% CI |  | OR<sup>a</sup> | 95% CI |  |
| Healthy (fifths/quintiles) |  |  |  |  |  |  |
| First |  | Referent |  |  | Referent |  |
| Second | 0.57 | 0.52 - 0.63 | *** | 0.87 | 0.74 - 1.03 |  |
| Third | 0.45 | 0.40 - 0.50 | *** | 0.88 | 0.72 - 1.07 |  |
| Fourth | 0.41 | 0.37 - 0.46 | *** | 0.88 | 0.70 - 1.12 |  |
| Fifth | 0.42 | 0.38 - 0.48 | *** | 0.97 | 0.73 - 1.29 |  |
| *Ordinal* | 0.80 | 0.78-0.82 | *** | 0.99 | 0.93 - 1.01 |  |
| *Less* healthy (fifths/quintiles) |  |  |  |  |  |  |
| First |  | Referent |  |  | Referent |  |
| Second | 1.24 | 1.11 - 1.37 |  | 1.07 | 0.91 - 1.27 |  |
| Third | 1.57 | 1.40 - 1.75 | *** | 1.07 | 0.87 - 1.31 |  |
| Fourth | 2.01 | 1.79 - 2.25 | *** | 1.03 | 0.81 - 1.31 |  |
| Fifth | 2.49 | 2.19 - 2.83 | *** | 1.02 | 0.76 - 1.36 |  |
| *Ordinal* | 1.27 | 1.23 - 1.30 | *** | 1.00 | 0.94 - 1.07 |  |
| Ratio (fifths/quintiles) |  |  |  |  |  |  |
| First |  | Referent |  |  | Referent |  |
| Second | 0.54 | 0.49 - 0.59 | *** | 1.00 | 0.90 - 1.11 |  |
| Third | 0.41 | 0.37 - 0.45 | *** | 1.01 | 0.90 - 1.12 |  |
| Fourth | 0.35 | 0.32 - 0.39 | *** | 1.05 | 0.94 - 1.17 |  |
| Fifth | 0.37 | 0.34 - 0.42 | *** | 1.26 | 1.13 - 1.40 | *** |
| *Ordinal* | 0.78 | 0.76 - 0.79 | *** | 1.05 | 1.03 - 1.08 | *** |

\*\*\* p<0.0001, \*\* p<0.001, \* p<0.01. Cumulative Time weighted Food Access scores (computed using the nearest distance to food outlets).
<sup>a</sup> Adjusted for age, year of entry, and exit from the study, individual-level race/ethnicity, reproductive factors (menopause, livebirths, breastfeeding, parity, age at menarche), tract-level SES factors (disadvantage, affluence, proportion of NH, White, Black, and Hispanic at tract level), family history of breast cancer and prior history of biopsy and, mutually adjusted for food access measures including distance to undetermined food outlets; models with healthy food scores are adjusted with scores for *less* healthy food and vice versa, except for the ratio measures that are implicitly adjusted.

*Driving distance:* Greater driving distance-based IDW scores for healthy food access were numerically but not statistically associated with a modest, monotonic decrease in incidence to a 20% reduced incidence in the highest compared to the lowest fifth (Table 3 and Figure 2).

There was no association of driving distance to the nearest healthy food outlet with incidence (Table 4 and Figure 3).

#### *Less* healthy food access

*Walking distance:* Incidence was 40% greater at the highest vs. the lowest quintile of walking distance-based IDW scores for *less* healthy food access (Table 3 and Figure 2).

Associations were stronger for walking distance to the *nearest less* healthy food outlet (Table 4 and Figure 3). Incidence increased monotonically to a 150% greater incidence in the highest vs. the lowest quintile of *less* healthy food access.

*Driving distance:* There was a slight trend of increased incidence with greater driving distance-based, IDW *less* healthy food access scores (Table 3 and Figure 2), and no association with driving distance to the nearest *less* healthy food outlet (Table 4 and Figure 3).

#### Ratio of healthy to *less* healthy food access

*Walking distance:* Relatively greater healthy food access demonstrated a U-shaped, association with breast cancer incidence, with ∼20% lower incidence in the second, third and fourth quintiles and returning to an incidence in the highest quintile similar to that for the lowest quintile (Table 3 and Figure 2). Relatively shorter distance to the nearest healthy food outlet was associated with a strong monotonic decrease in incidence, dropping to a nearly two-thirds reduced incidence in the highest vs. lowest quintile (Table 4 and Figure 3).

*Driving distance:* Relatively greater healthy food access was not associated with breast cancer incidence (Table 3 and Figure 2). Relatively shorter distance to the nearest healthy food outlet was associated with a 20% lower incidence when comparing the highest vs. the lowest quintile (Table 4 and Figure 3).

### By Menopausal Status

Among pre-menopausal women, breast cancer incidence was 45-60% lower for all categories of walking distance healthy food access IDW scores when compared to the lowest quintile of access. For postmenopausal women, breast cancer incidence declined in a U-shaped fashion and was 30-40% lower for all but the highest quintile of walking distance healthy food access scores when compared to the lowest quintile of access. Driving distance-based measures of healthy food access were generally not associated with incidence (Table S5). For *less* healthy food access, the highest walking distance scores for *less* healthy food access were associated with a 35-40% greater incidence when compared to the lowest quintile of access for both pre-menopausal and post-menopausal women. Greater walking distance-based relative access to healthy vs. *less* healthy food was generally associated with reduced incidence for both pre-menopausal and post- menopausal women. The corresponding driving distance-based measures, however, were (contrary to expectation) associated with greater incidence for pre-menopausal women and generally not associated with incidence for post-menopausal women (Table S5).

### Geospatial access to food outlets and incidence by tumor subtype

Regarding healthy food access, ER/PR negative breast cancer incidence was 30-40% lower for all levels of walking and driving distance-based healthy food access when compared to the lowest level of access, whereas associations were not apparent for ER/PR positive tumors.

Regarding *less* healthy food access, associations were variable and more modest in magnitude, regardless of tumor subtype. Relatively greater access to healthy vs. unhealthy food was generally not associated or inconsistently associated with incidence regardless of subtype (Table S6).

## DISCUSSION

This study examined the association between long-term geospatial access to both healthy and *le*ss healthy food outlets and breast cancer incidence among women in Metropolitan Chicago over nearly three decades. By leveraging longitudinal residential histories and geospatial techniques, our findings contribute to understanding how pre-diagnostic neighborhood food environments influence breast cancer risk. Consistent with our hypothesis, greater access to healthy food outlets was associated with lower breast cancer incidence, particularly for walking-based measures, emphasizing the role of walkable environments, which may increase opportunities for healthier food purchases and promote healthier behaviors.^33,36^ Studies have shown that living closer to supermarkets or fresh produce markets is associated with higher intake of fruits and vegetables, especially when the prices are affordable and stores are culturally appropriate.^33,37,38^ Conversely, greater access to *less* healthy food outlets was associated with increased breast cancer incidence, especially for walking-based measures, underscoring the detrimental effects of food deserts/swamps where the predominance of convenience stores and fast-food outlets may encourage unhealthy dietary patterns. These findings support existing evidence that proximity to *less* healthy food outlets may reinforce poor dietary patterns over time. In general, although proximity to healthy food outlets does not inherently ensure healthier dietary choices, it can facilitate greater exposure to nutritious options and increase the likelihood of engaging in health-promoting food purchasing behaviors. However, structural factors such as affordability, cultural preferences, and food literacy also play a critical role in shaping food consumption. Our findings underscore the need for food environment interventions that not only increase physical access to healthy options but also address structural and behavioral barriers to their use.

Greater access to healthy food was associated with reduced incidence for walking-based measures for pre-menopausal women. Driving-based measures showed a marginal reduction in incidence (only in lower quintiles), with the association plateauing for the highest healthy food access levels. Post-menopausal women also demonstrated lower breast cancer incidence for healthy food access, particularly for driving-based measures. In contrast, access to *less* healthy food was associated with greater incidence for both pre- and post-menopausal women, with pronounced effects observed for the highest quintiles of walking-based measures. Across tumor subtypes, healthy food access reduced incidence, especially for walking distances, with stronger effects for ER/PR-negative tumors.

A notable finding of this study is the U-shaped pattern in the association between healthy food access and breast cancer incidence. This trend highlights a threshold effect at the highest quintile of access, where a reduction in breast cancer incidence diminishes beyond the tipping point. One potential explanation could be that residents of urban areas with a high density of healthy food outlets are of higher socioeconomic status (SES) and have greater healthcare access; these, in turn, are associated with increased breast risk (higher SES) and increased detection (healthcare access). Residual confounding by these factors would be expected to attenuate the association of healthy food access with breast cancer incidence.

Prolonged exposure to healthy food environments may promote optimal dietary patterns, decreasing cancer risk via hormonal, inflammatory, and oxidative stress pathways, whereas prolonged exposure to *less* healthy food environments may have the opposite effect. Contrary to expectation, we found more consistent associations with respect to food access measures based on residential proximity to the nearest food outlet (both healthy and *less* healthy) than the more complex inverse-distance-based measures of density of food outlets. These findings suggest that residential proximity to at least one food outlet (both healthy and *less* healthy) may be a more important factor in shaping dietary behaviors and subsequent cancer incidence than the density of healthy or *less* healthy food outlets surrounding one’s residence. This aligns with prior literature showing mixed associations between food outlet density and diet or health outcomes, likely due to differences in how density is defined and measured across studies.^33,39,40^ In addition, we found more consistent associations with respect to food access measures based on walking distance to the nearest food outlet (both healthy and *less* healthy) than we did for driving distance to the nearest food outlet. It may be that in densely populated urban areas where many residents do not own a car, being able to walk to a nearby food outlet becomes more important than being able to drive to a possibly more distant food outlet. Although our study included both urban and suburban residences, we were unable to examine associations of food access with incidence stratified on urban vs. suburban residence because women’s residential histories typically included both urban and suburban residences.

Despite strengths such as incorporation of robust residential histories and geospatial analysis, this study has several limitations. The classification of food outlets may not fully encompass the diversity and complexity of dietary environments, with certain types (e.g., restaurants and delis) excluded. Dietary behaviors and affordability, an essential mediator between food access and health outcomes, could not be directly assessed. Although we included an array of hormonal, reproductive, and other patient factors along with census-tract disadvantage and affluence as model covariates, misclassification of these variables and absence of control for physical activity, alcohol intake, and other variables may lead to residual confounding. This study does not capture specific variables such as individual travel preferences, purchasing behaviors, or food environments beyond residential settings, such as workplaces or transit points. Moreover, the role of food delivery services or purchases made during commutes could not be captured in these analyses. While we incorporated up to 30 years of annual data on the food environment, residential histories were incomplete for many participants for at least a portion of this time period, and we assumed that these missing residential histories were missing at random (MAR), The reliance on inverse-distance weighting methods for food access scores assumes linear decay in influence, which may not fully capture the complexities of food access. Moreover, the generalizability of findings is limited to urban settings in the Chicago metropolitan area.

Expanding similar analyses to rural regions and other geographical areas would provide a more comprehensive understanding of the interplay between food environments and breast cancer risk across diverse populations. Future research could benefit from integrating consumer-level data, including dietary patterns and food purchasing habits, to strengthen the causal evidence for these associations.

In conclusion, this study advances our understanding of the impact of long-term geospatial access to food outlets on breast cancer incidence, with important implications for research and public health practice. Notably, it is the first to apply long-term, time-weighted geospatial food access metrics in breast cancer epidemiology. These findings extend prior research by emphasizing the relevance of long-term residential histories, rather than relying solely on diagnosis-year exposures, to understand the cumulative neighborhood impacts on breast cancer outcomes. Our results suggest that not just the presence of healthy food outlets, but also their proximity and walkability, may be critical. Accordingly, policy efforts should move beyond simply placing grocery stores in underserved areas and instead focus on improving walkable access to affordable, healthy food. This may include incentivizing grocery stores and farmers’ markets to offer competitively priced produce in high-need neighborhoods and adapting urban planning and zoning policies to both enhance walkability and limit the density of fast-food and convenience outlets in communities at elevated risk.

## Supporting information

All Supplementary data

## Data Availability

All data produced in the present study are available upon reasonable request to the authors

## REFERENCE

1. Giaquinto AN, Sung H, Miller KD, et al. Breast Cancer Statistics, 2022. CA Cancer J Clin. 2022;72(6):524–541. doi:10.3322/caac.21754

2. Patel KG, Borno HT, Seligman HK. Food insecurity screening: A missing piece in cancer management. Cancer. 2019;125(20):3494–3501. doi:10.1002/cncr.32291

3. Roy AM, George A, Attwood K, et al. Effect of neighborhood deprivation index on breast cancer survival in the United States. Breast Cancer Res Treat. 2023;202(1):139–153. doi:10.1007/s10549-023-07053-4

4. Cooksey-Stowers K, Schwartz MB, Brownell KD. Food swamps predict obesity rates better than food deserts in the United States. Int J Environ Res Public Health. 2017;14(11). doi:10.3390/ijerph14111366

5. Lathigara D, Kaushal D, Wilson RB. Molecular Mechanisms of Western Diet-Induced Obesity and Obesity-Related Carcinogenesis—A Narrative Review. Metabolites. 2023;13(5). doi:10.3390/metabo13050675

6. Seiler A, Chen MA, Brown RL, Fagundes CP. Obesity, Dietary Factors, Nutrition, and Breast Cancer Risk. Curr Breast Cancer Rep. 2018;10(1):14–27. doi:10.1007/s12609-018-0264-0

7. Ng WH, Abu Zaid Z, Mohd Yusof BN, Amin Nordin S, Lim PY. Association between dietary inflammatory index and body fat percentage among newly diagnosed breast cancer patients. Ann Med. 2023;55(2). doi:10.1080/07853890.2024.2303399

8. Munsell MF, Sprague BL, Berry DA, Chisholm G, Trentham-Dietz A. Body Mass Index and Breast Cancer Risk According to Postmenopausal Estrogen-Progestin Use and Hormone Receptor Status. Epidemiol Rev. 2014;36(1):114. doi:10.1093/EPIREV/MXT010

9. Renehan AG, Tyson M, Egger M, Heller RF, Zwahlen M. Body-mass index and incidence of cancer: a systematic review and meta-analysis of prospective observational studies. Lancet. 2008;371(9612):569-578. doi:10.1016/S0140-6736(08)60269-X

10. Schoemaker MJ, Nichols HB, Wright LB, et al. Association of Body Mass Index and Age with Subsequent Breast Cancer Risk in Premenopausal Women. JAMA Oncol. 2018;4(11). doi:10.1001/jamaoncol.2018.1771

11. Xiao Y, Xia J, Li L, et al. Associations between dietary patterns and the risk of breast cancer: A systematic review and meta-analysis of observational studies. Breast Cancer Research. 2019;21(1). doi:10.1186/s13058-019-1096-1

12. Castelló A, Rodríguez-Barranco M, Lope V, et al. High adherence to Western dietary pattern increases breast cancer risk (an EPIC-Spain study). Maturitas. 2024;179. doi:10.1016/j.maturitas.2023.107868

13. Cubbin C, Pedregon V, Egerter S, Braveman P. Where We Live Matters for Our Health: Neighborhoods and Health. Published online 2008. Accessed November 15, 2023. http://activateomaha.org/

14. Diez Roux A V., Mair C. Neighborhoods and health. Ann N Y Acad Sci. 2010;1186:125–145. doi:10.1111/J.1749-6632.2009.05333.X

15. Akinyemiju TF, Genkinger JM, Farhat M, Wilson A, Gary-Webb TL, Tehranifar P. Residential environment and breast cancer incidence and mortality: A systematic review and meta-analysis. BMC Cancer. 2015;15(1). doi:10.1186/s12885-015-1098-z

16. Dunn BK, Agurs-Collins T, Browne D, Lubet R, Johnson KA. Health disparities in breast cancer: Biology meets socioeconomic status. Breast Cancer Res Treat. 2010;121(2):281–292. doi:10.1007/s10549-010-0827-x

17. USDA ERS - Food Access Research Atlas. Accessed November 16, 2023. https://www.ers.usda.gov/data-products/food-access-research-atlas/

18. Araújo ML de, Mendonça R de D, Lopes Filho JD, Lopes ACS. Association between food insecurity and food intake. Nutrition. 2018;54:54–59. doi:10.1016/j.nut.2018.02.023

19. Pérez-escamilla R. Revista de Nutrição Food Insecurity Measurement and Indicators Indicadores e Medidas de Insegurança Alimentar. Vol 21.; 2008.

20. Kendall A, Olson CM, Frongillo EA. Relationship of Hunger and Food Insecurity to Food Availability and Consumption. J Am Diet Assoc. 1996;96(10):1019–1024. doi:10.1016/S0002-8223(96)00271-4

21. Lucan SC, Mitra N. The food environment and dietary intake: Demonstrating a method for GIS-mapping and policy-relevant research. Journal of Public Health (Germany). 2012;20(4):375–385. doi:10.1007/s10389-011-0470-y

22. Aretz B, Costa R, Doblhammer G, Janssen F. The association of unhealthy and healthy food store accessibility with obesity prevalence among adults in the Netherlands: A spatial analysis. SSM Popul Health. 2023;21. doi:10.1016/j.ssmph.2022.101332

23. Cobb LK, Appel LJ, Franco M, Jones-Smith JC, Nur A, Anderson CAM. The relationship of the local food environment with obesity: A systematic review of methods, study quality, and results. Obesity. 2015;23(7):1331–1344. doi:10.1002/oby.21118

24. Ojinnaka CO, Christ J, Bruening M. Is There a Relationship between County-Level Food Insecurity Rates and Breast Cancer Stage at Diagnosis? Nutr Cancer. 2022;74(4):1291–1298. doi:10.1080/01635581.2021.1952624

25. Namin S, Zhou Y, Neuner J, Beyer K. The role of residential history in cancer research: A scoping review. Soc Sci Med. 2021;270. doi:10.1016/j.socscimed.2020.113657

26. Stinchcomb DG, Roeser A. Westat An Employee-Owned Research Corporation ® 1600 Research. 2016;301:251–1500.

27. Dabbous FM, Dolecek TA, Berbaum ML, et al. Impact of a false-positive screening mammogram on subsequent screening behavior and stage at breast cancer diagnosis. Cancer Epidemiology Biomarkers and Prevention. 2017;26(3):397–403. doi:10.1158/1055-9965.EPI-16-0524

28. Rauscher GH, Dabbous F, Dolecek TA, et al. Absence of an anticipated racial disparity in interval breast cancer within a large health care organization. Ann Epidemiol. 2017;27(10):654–658. doi:10.1016/j.annepidem.2017.09.002

29. Powell LM, Han E, Zenk SN, et al. Field validation of secondary commercial data sources on the retail food outlet environment in the U.S. Health Place. 2011;17(5):1122–1131. doi:10.1016/J.HEALTHPLACE.2011.05.010

30. Miller C, Bodor JN, Rose D. Measuring the Food Environment: A Systematic Technique for Characterizing Food Stores Using Display Counts. J Environ Public Health. 2012;2012(1):707860. doi:10.1155/2012/707860

31. Hirsch JA, Zhao Y, Melly S, et al. National trends and disparities in retail food environments in the USA between 1990 and 2014. Public Health Nutr. 2023;26(5):1052. doi:10.1017/S1368980023000058

32. Ver Ploeg M, Breneman V, Farrigan T, et al. Access to affordable and nutritious food: measuring and understanding food deserts and their consequences: report to congress. Published online 2009.

33. Caspi CE, Sorensen G, Subramanian S V., Kawachi I. The local food environment and diet: A systematic review. Health Place. 2012;18(5):1172–1187. doi:10.1016/J.HEALTHPLACE.2012.05.006

34. Liese AD, Weis KE, Pluto D, Smith E, Lawson A. Food Store Types, Availability, and Cost of Foods in a Rural Environment. J Am Diet Assoc. 2007;107(11):1916–1923. doi:10.1016/j.jada.2007.08.012

35. Barber LE, Zirpoli GR, Cozier YC, et al. Neighborhood disadvantage and individual-level life stressors in relation to breast cancer incidence in US Black women. Breast Cancer Research. 2021;23(1):108. doi:10.1186/s13058-021-01483-y

36. Michele Ver Ploeg VBTFKHDHBHLMNTASRWKKCOAS and ET. Access to Affordable and Nutritious Food-Measuring and Understanding Food Deserts and Their Consequences: Report to Congress | Economic Research Service. Accessed July 8, 2025. https://www.ers.usda.gov/publications/pub-details?pubid=42729

37. Zenk SN, Lachance LL, Schulz AJ, Mentz G, Kannan S, Ridella W. Health promoting community design/nutrition: Neighborhood retail food environment and fruit and vegetable intake in a multiethnic urban population. American Journal of Health Promotion. 2009;23(4):255–264. doi:10.4278/AJHP.071204127,

38. Larson NI, Story MT, Nelson MC. Neighborhood Environments. Disparities in Access to Healthy Foods in the U.S. Am J Prev Med. 2009;36(1). doi:10.1016/j.amepre.2008.09.025

39. Pineda E, Stockton J, Scholes S, Lassale C, Mindell JS. Food environment and obesity: a systematic review and meta-analysis. BMJ Nutr Prev Health. 2024;7(1):204–211. doi:10.1136/BMJNPH-2023-000663

40. Fiechtner L, Block J, Duncan DT, et al. Proximity to Supermarkets Associated with Higher Body Mass Index among Overweight and Obese Preschool-Age Children. Prev Med (Baltim). 2012;56(0):10.1016/j.ypmed.2012.11.023. doi:10.1016/J.YPMED.2012.11.023

