## Supplementary material for "Longer-Term Geospatial Food Access and the Incidence of Breast Cancer in Metropolitan Chicago": All Supplementary data

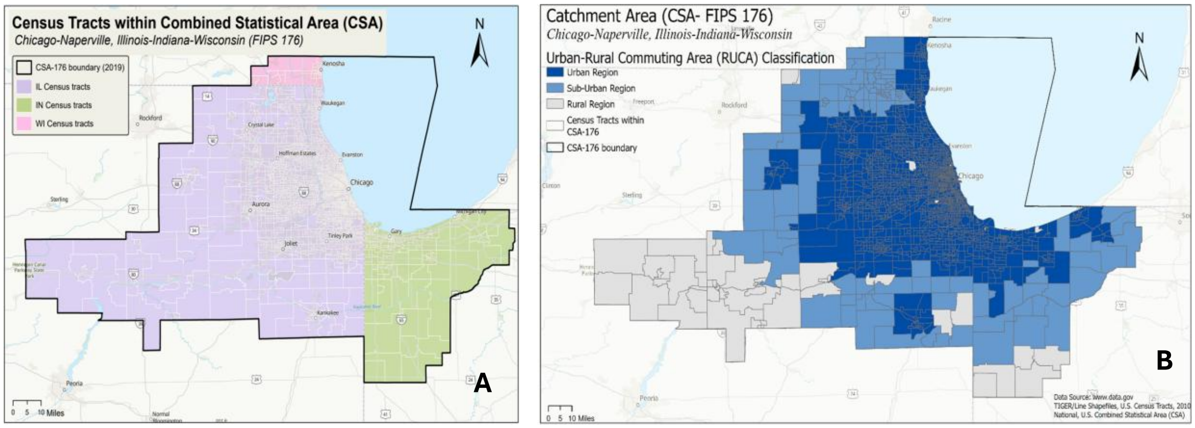


**Figure S1. Catchment area (CSA-176), as defined by the U.S. Census Bureau**. A. Catchment area covering 19 counties: 13 in Illinois (Bureau, Cook, DeKalb, DuPage, Grundy, Kane, Kankakee, Kendall, Lake, LaSalle, McHenry, Putnam, Will), 5 in Indiana (Jasper, Lake, LaPorte), and 1 in Wisconsin (Kenosha). The map displays all census tracts within this area. B. Urban-rural classification of the catchment area using RUCA codes. Over 90% of the residences used in this study were predominantly urban or suburban (point addresses- not shown).

Table S1. Categorizing food outlets in the NETS dataset 1990-2019 (N=310,981)

| Identified using | Healthy<br>N=131,657<br>(42.34%) | *Less* healthy<br>N=108837<br>(35.00%) | Undetermined<br>N=70487<br>(22.67%) |
|---|---|---|---|
| SIC, industry type, company and tradenames | Grocery stores (chain, independent), Supermarkets (chain, independent, small, or large) and Fruit and vegetable markets/stands | Convenience stores (chain, independent), Gasoline stations with convenience stores, Fast-food places and stands (carry out, drive-in, chain, independent) | Cafe, Restaurants, Delicatessen stores, food bars, grill (eating places), hamburger/hotdog stands, sandwich shops and snack bars |

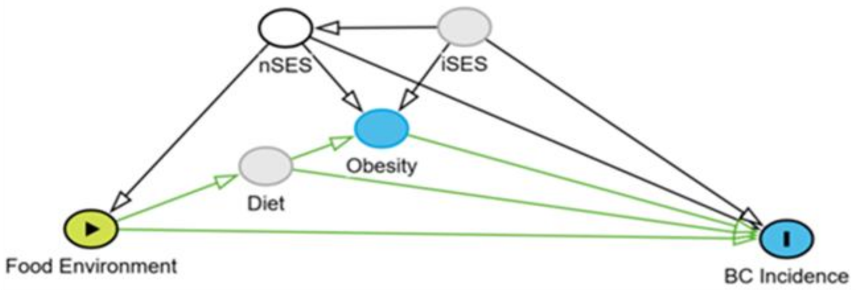


**Figure S2. Directed Acyclic Graph (DAG) for association between geospatial food access and incidence of breast cancer.** Food environment is the exposure variable that includes walking and driving distance for access to healthy and less healthy food outlets and the relative access (ratio of healthy to less healthy food outlets). BC (Breast cancer) Incidence is the outcome variable, Obesity is measured using BMI (Body-Mass-Index; continuous scale), Diet: Personal diet choices and consumption are unobserved, nSES: neighborhood socio-economic factors, measured using disadvantage and affluence, and tract level proportion of White, NH, Black NH, and Hispanic race/ethnicity, iSES: individual level socio-economic status is unobserved. Age, and year of entry and exit in the study (not shown in the figure) are included a-priori. Race/ethnicity, family history of breast cancer, and reproductive factors (breastfeeding, parity, livebirths, age at menarche) are not shown but are controlled for in all models.

| Table S2: Summary statistics of food access scores (1990-2019) | | | | | | | |
|---|---|---|---|---|---|---|---|
| ***Mode of Access: Walking*** | | | | | | | |
| Geospatial Food Access Measure | N | Mean | SD | Median | Variance | Skewness | Kurtosis |
| Healthy food (IDW) | 29,216 | 1.81 | 0.84 | 1.69 | 0.71 | 0.92 | 4.56 |
| *Less* healthy food (IDW) | 29,216 | 8.93 | 2.36 | 8.81 | 5.57 | 0.23 | 3.91 |
| Relative scale (IDW) | 29,212 | 0.21 | 0.12 | 0.19 | 0.02 | 5.64 | 105.47 |
| Healthy food (Nearest distance) | 28, 840 | 0.52 | 0.15 | 0.51 | 0.02 | 0.85 | 5.93 |
| *Less* healthy food (Nearest distance) | 29,194 | 0.48 | 0.12 | 0.46 | 0.02 | 1.38 | 8.89 |
| Relative scale (Nearest distance) | 28,810 | 1.11 | 0.33 | 1.08 | 0.11 | 2.85 | 47.14 |
| ***Mode of Access: Driving*** | | | | | | | |
| Geospatial Food Access Measure | N | Mean | SD | Median | Variance | Skewness | Kurtosis |
| Healthy food (IDW) | 29,248 | 3.24 | 2.54 | 2.56 | 6.45 | 2.13 | 8.98 |
| *Less* healthy food (IDW) | 29,248 | 14.84 | 8.43 | 13.1 | 71.12 | 1.86 | 8.98 |
| Relative scale (IDW) | 29,248 | 0.25 | 0.86 | 0.21 | 0.74 | 159.42 | 26624.64 |
| Healthy food (Nearest distance) | 29,247 | 0.45 | 0.42 | 0.29 | 0.18 | 2.15 | 9.82 |
| *Less* healthy food (Nearest distance) | 29,248 | 0.25 | 0.38 | 0.22 | 0.15 | 2.97 | 17.77 |
| Relative scale (Nearest distance) | 29,246 | 1.41 | 0.33 | 1.39 | 0.11 | 1.26 | 10.3 |

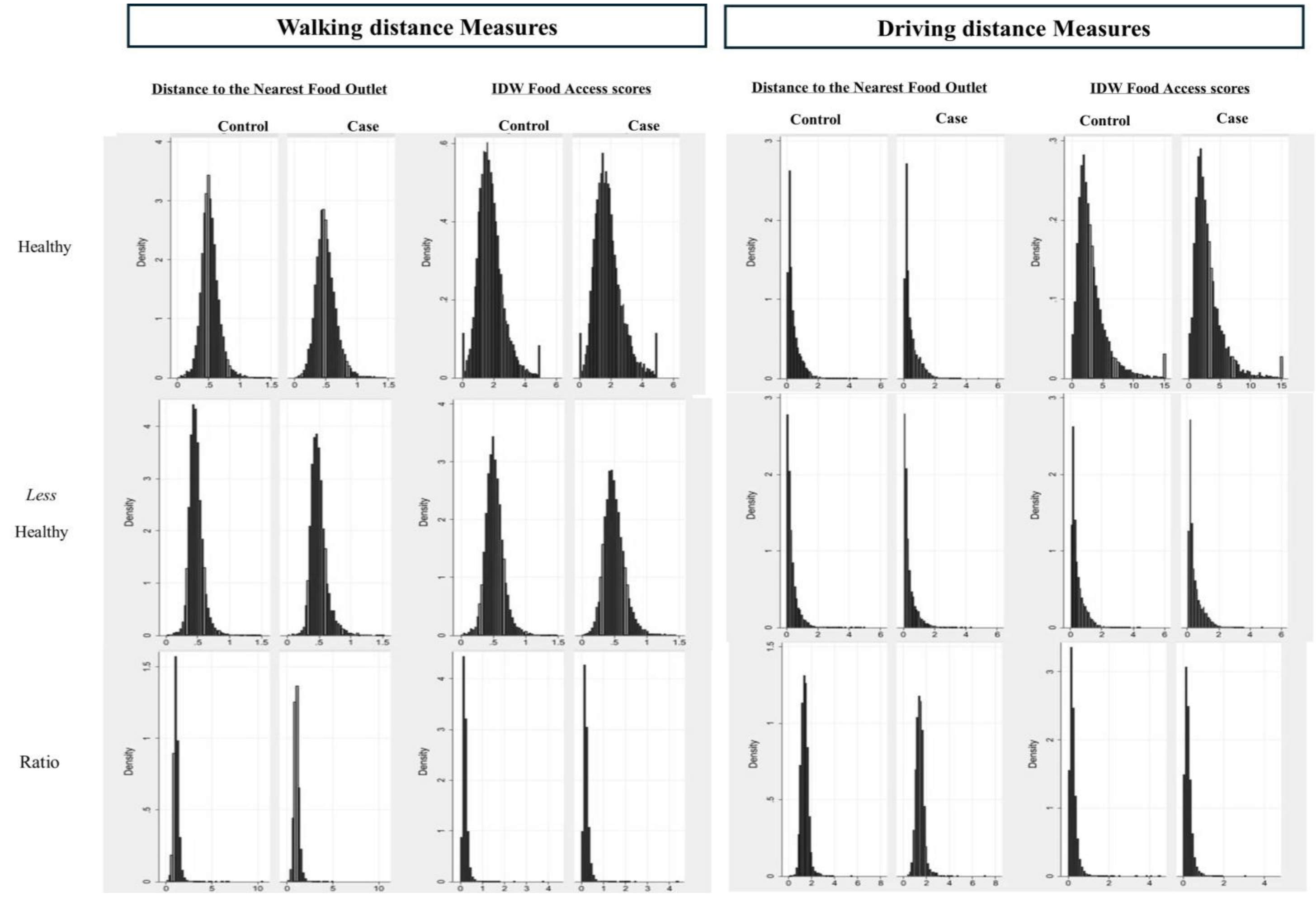


**Figure S3.** Distribution Food Access scores for both walking and driving as the mode of access for cases vs. controls depicted by histograms; nearest distance to healthy food outlets for walking (A) and driving (C); nearest distance to less healthy food outlets for walking (E) and driving (G); nearest distance measures for ratio of healthy to less healthy food for walking (I) and driving (K); IDW access to healthy food outlets for walking (B) and driving (D); IDW access to *less* healthy food outlets for walking (F) and driving (H); IDW access to ratio of healthy to *less* healthy food for walking (J) and driving (L). For IDW ratio measures, one observation with score=143.5 was excluded (data not shown).

Table S3: Crude Distribution of 30-year (1990-2019) inverse distance time-weighted (IDW) walking and driving distance-based food access scores in breast cancer case-control study

| | Case; n=7376 | | Control; n=21,900 | |
|---|---|---|---|---|
| | **n** | **%** | **n** | **%** |
| *Walking distance scores* | | | | |
| Healthy (fifths/quintiles) | | | | |
| First | 1586 | 21% | 4258 | 19% |
| Second | 1406 | 19% | 4437 | 20% |
| Third | 1378 | 19% | 4465 | 20% |
| Fourth | 1391 | 19% | 4452 | 20% |
| Fifth | 1618 | 22% | 4225 | 19% |
| Missing | 17 | | 63 | |
| *Less* Healthy (fifths/quintiles) | | | | |
| First | 1549 | 21% | 4295 | 20% |
| Second | 1317 | 18% | 4526 | 21% |
| Third | 1361 | 18% | 4482 | 21% |
| Fourth | 1446 | 20% | 4397 | 20% |
| Fifth | 1706 | 23% | 4137 | 19% |
| Missing | 17 | | 63 | |
| Ratio (fifths/quintiles) | | | | |
| First | 1561 | 21% | 4282 | 20% |
| Second | 1402 | 19% | 4440 | 20% |
| Third | 1420 | 19% | 4423 | 20% |
| Fourth | 1396 | 19% | 4446 | 20% |
| Fifth | 1598 | 22% | 4244 | 19% |
| Missing | 19 | | 65 | |
| *Driving distance measures* | | | | |
| Healthy (fifths/quintiles) | | | | |
| First | 1437 | 19% | 4413 | 20% |
| Second | 1516 | 21% | 4334 | 20% |
| Third | 1497 | 20% | 4352 | 20% |
| Fourth | 1481 | 20% | 4369 | 20% |
| Fifth | 1451 | 20% | 4398 | 20% |
| Missing | 14 | | 34 | |
| *Less* Healthy (fifths/quintiles) | | | | |
| First | 1576 | 21% | 4274 | 20% |
| Second | 1486 | 20% | 4364 | 20% |
| Third | 1482 | 20% | 4367 | 20% |
| Fourth | 1451 | 20% | 4399 | 20% |
| Fifth | 1387 | 19% | 4462 | 20% |
| Missing | 14 | | 34 | |
| Ratio (fifths/quintiles) | | | | |
| First | 1394 | 19% | 4456 | 20% |
| Second | 1362 | 18% | 4488 | 21% |
| Third | 1458 | 20% | 4391 | 20% |
| Fourth | 1534 | 21% | 4316 | 20% |
| Fifth | 1634 | 22% | 4215 | 19% |
| Missing | 14 | | 34 | |

Table S4: Crude Distribution of 30-year (1990-2019) time-weighted nearest walking and driving distance-based food access scores in breast cancer case-control study

| | Case (n=7396) | | Control (n=21,900) | |
|---|---|---|---|---|
| | **n** | **%** | **n** | **%** |
| *Walking distance scores* | | | | |
| Healthy (fifths/quintiles) | | | | |
| First | 1968 | 27% | 3800 | 17% |
| Second | 1424 | 19% | 4344 | 20% |
| Third | 1209 | 16% | 4559 | 21% |
| Fourth | 1205 | 16% | 4563 | 21% |
| Fifth | 1471 | 20% | 4297 | 20% |
| Missing | 119 | | 337 | |
| *Less* Healthy (fifths/quintiles) | | | | |
| First | 1299 | 18% | 4540 | 21% |
| Second | 1296 | 18% | 4543 | 21% |
| Third | 1366 | 18% | 4473 | 20% |
| Fourth | 1533 | 21% | 4306 | 20% |
| Fifth | 1876 | 25% | 3962 | 18% |
| Missing | 26 | | 76 | |
| Ratio (fifths/quintiles) | | | | |
| First | 2208 | 30% | 3559 | 16% |
| Second | 1558 | 21% | 4208 | 19% |
| Third | 1278 | 17% | 4488 | 20% |
| Fourth | 1128 | 15% | 4638 | 21% |
| Fifth | 1102 | 15% | 4664 | 21% |
| Missing | 122 | | 343 | |
| *Driving distance measures* | | | | |
| Healthy (fifths/quintiles) | | | | |
| First | 1426 | 19% | 4424 | 20% |
| Second | 1518 | 21% | 4331 | 20% |
| Third | 1466 | 20% | 4384 | 20% |
| Fourth | 1377 | 19% | 4472 | 20% |
| Fifth | 1595 | 22% | 4254 | 19% |
| Missing | 14 | | 35 | |
| *Less* Healthy (fifths/quintiles) | | | | |
| First | 1406 | 19% | 4444 | 20% |
| Second | 1562 | 21% | 4288 | 20% |
| Third | 1499 | 20% | 4350 | 20% |
| Fourth | 1309 | 18% | 4541 | 21% |
| Fifth | 1606 | 22% | 4243 | 19% |
| Missing | 14 | | 34 | |
| Ratio (fifths/quintiles) | | | | |
| First | 1500 | 20% | 4350 | 20% |
| Second | 1400 | 19% | 4449 | 20% |
| Third | 1354 | 18% | 4495 | 21% |
| Fourth | 1434 | 19% | 4415 | 20% |
| Fifth | 1694 | 23% | 4155 | 19% |
| Missing | 14 | | 36 | |

Table S5. Associations of healthy, *less* healthy, and ratio of healthy to *less* healthy food access with breast cancer incidence (both walking and driving distance-based measures), separately for pre-menopausal and post-menopausal patients.

| | Pre-menopausal | | | | | | Post-menopausal | | | | | |
|---|---|---|---|---|---|---|---|---|---|---|---|---|
| | Walking | | | Driving | | | Walking | | | Driving | | |
| | OR [a] | 95% CI | | OR [a] | 95% CI | | OR [a] | 95% CI | | OR [a] | 95% CI | |
| Healthy (fifths/quintiles) | | | | | | | | | | | | |
| First | Referent | | | Referent | | | Referent | | | Referent | | |
| Second | 0.53 | 0.39 - 0.73 | ** | 0.97 | 0.72 - 1.32 | | 0.77 | 0.67 - 0.88 | ** | 1.00 | 0.88 - 1.14 | |
| Third | 0.47 | 0.31 - 0.71 | ** | 1.06 | 0.71 - 1.58 | | 0.78 | 0.65 - 0.95 | * | 0.90 | 0.75 - 1.08 | |
| Fourth | 0.39 | 0.23 - 0.68 | ** | 0.96 | 0.57 - 1.60 | | 0.77 | 0.60 - 0.98 | * | 0.87 | 0.68 - 1.09 | |
| Fifth | 0.55 | 0.29 - 1.07 | * | 1.15 | 0.61 - 2.17 | | 0.98 | 0.72 - 1.34 | | 0.79 | 0.58 - 1.06 | |
| *Ordinal* | 0.92 | 0.79 - 1.08 | | 1.03 | 0.88 - 1.21 | | 1.01 | 0.94 - 1.10 | | 0.94 | 0.87 - 1.01 | * |
| *Less* healthy (fifths/quintiles) | | | | | | | | | | | | |
| First | Referent | | | Referent | | | Referent | | | Referent | | |
| Second | 0.87 | 0.66 - 1.14 | | 0.79 | 0.60 - 1.04 | * | 0.84 | 0.74 - 0.94 | ** | 1.02 | 0.90 - 1.15 | |
| Third | 0.88 | 0.67 - 1.15 | | 0.82 | 0.59 - 1.15 | | 0.88 | 0.79 - 0.99 | * | 1.12 | 0.97 - 1.30 | |
| Fourth | 0.88 | 0.67 - 1.14 | | 0.72 | 0.48 - 1.08 | | 1.02 | 0.91 - 1.15 | | 1.21 | 1.01 - 1.45 | * |
| Fifth | 1.35 | 1.05 - 1.74 | * | 0.74 | 0.45 - 1.21 | | 1.41 | 1.26 - 1.59 | ** | 1.24 | 1.00 - 1.54 | * |
| *Ordinal* | 1.08 | 1.02 - 1.15 | ** | 0.94 | 0.84 - 1.06 | | 1.10 | 1.07 - 1.13 | ** | 1.06 | 1.01 - 1.12 | * |
| Ratio (fifths/quintiles) | | | | | | | | | | | | |
| First | Referent | | | Referent | | | Referent | | | Referent | | |
| Second | 0.60 | 0.47 - 0.77 | ** | 1.17 | 0.87 - 1.57 | | 0.82 | 0.73 - 0.91 | ** | 0.88 | 0.78 - 1.00 | * |
| Third | 0.65 | 0.51 - 0.83 | ** | 1.43 | 1.00 - 2.04 | * | 0.81 | 0.72 - 0.90 | ** | 0.94 | 0.79 - 1.10 | |
| Fourth | 0.67 | 0.53 - 0.85 | ** | 1.64 | 1.05 - 2.56 | * | 0.82 | 0.74 - 0.92 | ** | 0.93 | 0.75 - 1.14 | |
| Fifth | 0.91 | 0.72 - 1.14 | | 1.87 | 1.08 - 3.26 | * | 1.03 | 0.92 - 1.15 | | 0.95 | 0.73 - 1.23 | |
| *Ordinal* | 0.99 | 0.94 - 1.04 | | 1.17 | 1.02 - 1.34 | * | 1.01 | 0.98 - 1.03 | | 1.00 | 0.93 - 1.06 | |

*** $p<0.0001$, ** $p<0.001$, * $p<0.01$ ^Cumulative Time weighted Food Access scores (computed using the Inverse Distance weighting method).

a Adjusted for age, year of entry, and exit from the study, individual-level race/ethnicity, reproductive factors (menopause, livebirths, breastfeeding, parity, age at menarche), tract-level SES (disadvantage, affluence, proportion of NH, White, Black, and Hispanic at tract level), family history of breast cancer and prior history of biopsy and, mutually adjusted for food access scores; models with healthy food scores are adjusted with scores for *less* healthy food and vice versa, except for the ratio measures that are implicitly adjusted.

Table S6. Associations of healthy and *less* healthy food access with breast cancer incidence for walking and driving distances, and on absolute and relative scales, separately for ER/PR negative, ER/PR positive breast cancer patients compared to all controls.

| | ER/PR Negative | | | | ER/PR Positive | | | |
|---|---|---|---|---|---|---|---|---|
| | Walking | | Driving | | Walking | | Driving | |
| | OR [a] | 95% CI | OR [a] | 95% CI | OR [a] | 95% CI | OR [a] | 95% CI |
| Healthy (fifths/quintiles) | | | | | | | | |
| First | Referent | | Referent | | Referent | | Referent | |
| Second | 0.62 | 0.38-1.00 | 0.67 | 0.60 - 1.07 | 1.04 | 0.70-1.56 | 1.05 | 0.83 - 1.34 |
| Third | 0.60 | 0.34-1.06 | 0.53 | 0.49 - 1.05 | 0.95 | 0.59-1.53 | 0.92 | 0.66 - 1.28 |
| Fourth | 0.60 | 0.32-1.13 | 0.54 | 0.39- 1.07 | 1.01 | 0.59-1.72 | 0.95 | 0.62 - 1.45 |
| Fifth | 0.61 | 0.30-1.23 | 0.59 | 0.36 - 1.18 | 0.98 | 0.55-1.75 | 1.02 | 0.60 - 1.75 |
| *Ordinal* | 0.98 | 0.84 - 1.03 | 0.91 | 0.78 - 1.07 | 0.97 | 0.84 - 1.02 | 1.00 | 0.88 - 1.14 |
| *Less* healthy (fifths/quintiles) | | | | | | | | |
| First | Referent | | Referent | | Referent | | Referent | |
| Second | 0.94 | 0.73-1.21 | 1.01 | 0.78- 1.31 | 1.15 | 0.94-1.41 | 1.10 | 0.89 - 1.34 |
| Third | 1.07 | 0.84-1.36 | 1.09 | 0.85 - 1.41 | 1.14 | 0.93-1.39 | 1.11 | 0.80 - 1.22 |
| Fourth | 0.83 | 0.65-1.07 | 1.07 | 0.82 - 1.38 | 1.24 | 1.01-1.52 | 1.14 | 0.85 - 1.29 |
| Fifth | 0.93 | 0.74-1.19 | 0.99 | 0.71 - 1.24 | 1.21 | 0.99-1.47 | 1.18 | 0.86 - 1.32 |
| *Ordinal* | 0.98 | 0.93 - 1.03 | 0.99 | 0.93 - 1.06 | 1.05 | 0.99 - 1.09 | 0.98 | 0.92 - 1.03 |
| Ratio (fifths/quintiles) | | | | | | | | |
| First | Referent | | Referent | | Referent | | Referent | |
| Second | 0.83 | 0.66-1.05 | 1.01 | 0.89 - 1.21 | 1.11 | 0.92-1.35 | 0.91 | 0.72 - 1.14 |
| Third | 0.86 | 0.68-1.09 | 1.08 | 0.86 - 1.36 | 1.03 | 0.85-1.25 | 0.8 | 0.62 - 1.04 |
| Fourth | 0.92 | 0.73-1.17 | 1.08 | 0.85 - 1.36 | 0.99 | 0.82-1.21 | 0.95 | 0.72 - 1.24 |
| Fifth | 0.96 | 0.77-1.21 | 1.00 | 0.91 - 1.26 | 0.89 | 0.74-1.08 | 0.82 | 0.58 - 1.10 |
| *Ordinal* | 1.00 | 0.89 - 1.06 | 1.02 | 0.97 - 1.08 | 0.96 | 0.92 - 1.00 | 0.92 | 0.83 - 1.04 |

*** $p<0.0001$, ** $p<0.001$, * $p<0.01$ ^ Cumulative Time weighted Food Access scores (computed using the Inverse Distance weighting method). a Adjusted for age, year of entry, and exit from the study, individual-level race/ethnicity, reproductive factors (menopause, livebirths, breastfeeding, parity, age at menarche), tract-level SES (disadvantage, affluence, proportion of NH, White, Black, and Hispanic at tract level), family history of breast cancer and prior history of biopsy and, mutually adjusted for food access scores; models with healthy food scores are adjusted with scores for *less* healthy food and vice versa, except for the ratio measures that are implicitly adjusted.
